# Cognitive status and rehabilitation outcomes of patients in acute rehabilitation post Covid-19

**DOI:** 10.1101/2022.09.10.22279806

**Authors:** Madli Vahtra, Kyle Fahey, Aaron Malina, Sean Dreyer, Elliot Roth, Jordan Grafman, Prakash Jayabalan, Shira Cohen-Zimerman

## Abstract

**Objective:** This study aims to 1) characterize cognitive functioning in patients admitted for inpatient rehabilitation due to Covid-19 diagnosis and 2) examine how cognitive status at admission is associated with rehabilitation outcomes.

**Design:** Retrospective chart review.

**Setting:** An inpatient rehabilitation center located in Chicago, Illinois.

**Participants:** 80 participants in acute rehabilitation due to Covid-19 disease

**Intervention:** Not applicable.

**Main Outcome Measures:** Cognitive functioning as measured by the Montreal Cognitive Assessment (MoCA) and rehabilitation outcomes as measured by Functional Index Measure (FIM) and Section GG items for self-care and mobility (GG-SC and GG-M respectively).

**Results:** On average, our sample presented with mild cognitive impairment as assessed by the (MoCA). The most significant deficits were demonstrated in executive function, attention, language, and delayed free recall measures. Higher levels of overall cognitive function were associated with higher cognitive measures of rehabilitation outcomes. Weaker associations were observed with outcome measures of self-care and motor functioning.

**Conclusion:** Cognitive impairments are common in patients in acute rehabilitation due to Covid-19 and cognitive performance may help predict rehabilitation outcomes.

## Introduction

As the pandemic caused by the SARS-CoV-2 virus enters its third year, we are witnessing record breaking numbers of people diagnosed with Covid-19, with more than 382 million infections worldwide to date^1^. Many patients with severe Covid-19 disease experience a variety of problems with normal functioning and require rehabilitation services to overcome these problems^2–4^.

Covid-19 most commonly causes respiratory symptoms^5,6^. However, it has also been shown to impact the central and peripheral nervous systems^7–9^. Specifically, there is data to support that Covid-19 is associated with a unique pattern of cognitive impairments, namely in attention, executive function, and working memory. One study of 35 patients with Covid-19 infection found frequent impairments within memory, semantic fluency, mental flexibility, and phonetic fluency domains^10^. Another study analyzed cognitive test data collected from 84,285 participants with Covid-19 disease of varying severity^11^. The authors found deficits in global cognitive function, semantic problem solving, visual selective attention and executive function. More recent studies continue to show consistent findings. One reported significant cognitive decline in 93 patients with mild Covid-19 disease who did not require hospitalization or oxygen therapy^12^. Another, larger study consisting of 740 participants found significant cognitive deficits particularly in executive function, memory, and recall among hospitalized participants compared to those treated outpatient or in the Emergency Department^13^. While our understanding of the cognitive profile of people recovering from Covid-19 continues to expand, it appears that little attention has been given to the growing number of patients requiring acute rehabilitation following a Covid-19 diagnosis. Moreover, given that cognitive status is known to be associated with rehabilitation outcomes in older patients and those who suffer strokes^14–16^, it is relevant to test whether cognitive status of patients in acute rehabilitation post Covid-19 can predict rehabilitation outcomes. Yet, there are curently no peer-revied studies that address this question.

The current study aims to: (1) identify impaired cognitive domains in a sample of patients in acute rehabilitation post-covid-19 and (2) examine the association between cognitive functioning as assessed on admission and rehabilitation outcomes. To address these aims, we studied a group of 80 patients in an acute, inpatient rehabilitation center who were admitted due to a prior Covid-19 diagnosis. Based on existing literature^10–13^, we hypothesized that this group of patients would show cognitive impairments in domains of attention, executive function, and working memory. We also hypothesized that cognitive status would be associated with rehabilitation outcomes, including negatively associated with length of hospital stay and positively associated with functional outcomes as measured by the Functional Independence Measure (FIM) and Section GG items for self-care and mobility (GG-SC and GG-M respectively).

## Methods

### Participants

Participants were all adult patients who were admitted to an inpatient physical medicine and rehabilitation center in Chicago, Illinois following discharge from a general hospital where they were treated for acute Covid-19 between April 2020 and June 2020. Inclusion criteria included confirmed Covid-19 as the primary reason for hospitalization. All participants experienced some form of respiratory failure associated with their Covid-19 diagnosis. Individuals who were admitted for reasons other than Covid-19 and were later diagnosed with Covid-19 while in rehabilitation were excluded from this study. Formal Covid-19 diagnosis was established through a PCR test done at the general hospital where the patient was initially admitted prior to admission to the rehabilitation center. Given that all patients in our sample were hospitalized due to their Covid-19 symptoms and required rehabilitation following their hospitalization, they were all considered to be severe cases of Covid-19. The study was approved by the Northwestern University’s institutional review board (IRB).

### Data Collection

Electronic health records were retrospectively reviewed for all patients and the following information was obtained: age, sex, gender, race, years of education, and self-reported depression and anxiety levels (which were measured using the Patient Health Questionnaire (PHQ-9) and the General Anxiety Disorder Scale (GAD-7) respectively). Rehabilitation outcomes including duration of acute rehabilitation, GG-SC, GG-M, and FIM scores were collected at the time of discharge from rehabilitation.

### Outcome Variables

#### Cognitive functioning

Overall cognitive performance was evaluated using the Montreal Cognitive Assessment (MoCA). The MoCA is a 30-point cognitive screening tool used to estimate cognitive function and determine the need for patients to undergo further diagnostic testing. The tool evaluates the following cognitive domains and provides a sub-score for each: executive function (scores range from 0-5), naming (0-3), attention (0-6), language (0-3), abstraction (0-2), delayed recall (0-5), and orientation (0-6). A total MoCA score of 26 or higher indicates no cognitive impairment while scores between 18 and 25 indicate mild impairment, 10-17 indicate moderate impairment, and less than 10 indicate severe impairment.^17^ Of note, subscore values have been demonstrated to align with scores on more detailed neuropsychological tests, confirming their validity to stand as lone values outside of a total MoCA score.^18^

#### Rehabilitation outcomes

The Functional Independence Measure (FIM) was used to measure rehabilitation outcomes in cognition. The FIM is an 18-item, 7-level scale that evaluates motor and cognitive performance. Thirteen of the items evaluate motor function, 5 of the items evaluate cognitive function. For our study, data was collected only for the cognitive scale which included participant comprehension, expression, social interaction, problem solving, and memory. The FIM scoring system is as follows: 1= total assistance, 2=maximal assistance, 3=moderate assistance, 4=minimal assistance, 5= supervision, 6=modified independence, 7=complete independence^19^.

We used Section GG items for self-care (GG-SC) and mobility (GG-M) to evaluate functional status at the time of discharge. Section GG is a set of standardized assessment elements used to measure functional status of patients in inpatient rehabilitation facilities. The GG-SC includes 7 items, and the GG-M includes 17 items. We evaluated all GG-SC items and 8 of the 17 GG-M items. Each item in both scales is scored on a scale from 1 to 6 as follows: 1=dependent, 2=substantial/maximal assistance, 3=partial/moderate assistance, 4=supervision or touching assistance, 5= set up or cleanup assistance, 6= independence^20^. We also evaluated rehabilitation outcomes in terms of the duration of rehabilitation (in days) for each participant.

### Statistical Analysis

Statistical analyses were performed using JASP software (Version 0.11.1) on a macOS (Version 11.2.1). We performed classical hypothesis tests, including Spearman’s rho and Mann-Whitney U tests. Non-parametric testing was used because our data did not follow a normal distribution.

In addition, we performed Bayesian hypothesis tests, as the practice of wholly relying on p-values for statistical evidence has been repeatedly criticized. Benefits of including Bayesian analysis include the ability to account for unobservable variables and to make a more direct statement about the probability of a true hypothesis^21–25^. A BF_10_ of less than 0.33 and greater than 3 indicates moderate to strong evidence that the relationship is significant, with the strength increasing as the value moves further away from one. Values ranging from 0.33-3 indicates anecdotal evidence for a relationship, with a score of 1 indicating no evidence^26^.

## Results

### Overall Sample Characteristics

A total of 80 participants (31 female, 49 male) were included in the study. Patients ranged in age from 23 to 85 (Mean=61.89, SD=13.3). 33.75% were Caucasian, 40% were Black, and 26.25% were listed as another race or unknown. Participants’ years of education ranged between 2 and 21 years (Mean=14, SD=3.6). On average, they spent 21.6 days as inpatients in rehabilitation (range: 3-52 days). Mean and median depression scores amongst all 80 participants was 4.1 (SD=3.6) and 3 respectively, indicating minimal/normal levels of depression. Mean and median anxiety scores were 2.6 (SD=3.2) and 2 respectively, indicating minimal/normal anxiety.

### Cognitive Function

Out of the 80 study participants, 58 completed the entire MoCA and therefore, obtained a total score. Between 59 and 76 patients completed at least one subsection of the MoCA, as noted in Table 1. Participants who did not complete the entire MoCA were unable to do so due to motor or sensory symptoms. Of all participants who completed the entire MoCA, 41 (70.7%) of them demonstrated some form of cognitive impairment (total score <26). Seventeen showed no impairment (29.3%), 32 showed mild impairment (55.2%) 9 showed moderate impairment (15.5%) and 0 showed severe impairment. A distribution of total MoCA scores amongst these 58 participants can be found in Figure 1.

**Table 1:** Total Montral Cognitive Assessment (MoCA)and MoCA subscores are presented both as a whole group and separately for higher functioning patients (total MoCA score ≥26) and lower functioning (total MoCA score <26). Statistically significant findings are in bold (p<0.01). SD= standard deviation, U= Mann-Whitney U value, BF_10_= Bayesian factor. *There was no variance in Orientation scale in the high functioning group and therefore a value could not be obtained.

| MOCA Scores | N | Overall Mean (SD) | Median | Mean (SD): Higher Functioning | Mean (SD): Lower Functioning | U, P-value and BF<sub>10</sub> |
| --- | --- | --- | --- | --- | --- | --- |
| Total | 58 | 22.2 (4.4) | 23 | 27.2 (1.0) | 20.1 (3.4) | **U=697.0 (p< 0.00), BF<sub>10</sub>=957.0** |
| Executive Function | 59 | 3.5 (1.3) | 4 | 4.6 (0.7) | 3.1 (1.2) | **U=601.5 (p<0.001), BF<sub>10</sub>=35.2** |
| Naming | 73 | 2.6 (0.7) | 3 | 2.9 (0.3) | 2.6 (0.7) | U=440 (p=0.037), BF<sub>10</sub>= 0.6 |
| Attention | 69 | 4.6 (1.5) | 5 | 5.6 (0.5) | 4.2 (1.6) | **U=551.5 (p<0.001), BF<sub>10</sub>=6.8** |
| Language | 73 | 2.4 (0.7) | 2 | 5.8 (0.4) | 4.9 (1.0) | **U=558.5 (p<0.001), BF<sub>10</sub>= 7.9** |
| Abstraction | 74 | 1.3 (0.7) | 1 | 1.6 (0.5) | 1.2 (0.7) | U=433.5 (p=0.112), BF<sub>10</sub>= 0.5 |
| Memory | 76 | 1.8 (1.6) | 2 | 3.4 (1.0) | 1.3 (1.3) | **U=618.0 (p<0.001), BF<sub>10</sub>= 42.4** |
| Orientation | 75 | 5.6 (1.1) | 6 | 6.0 (0) | 5.4 (1.3) | N/A* |

**Figure 1:**
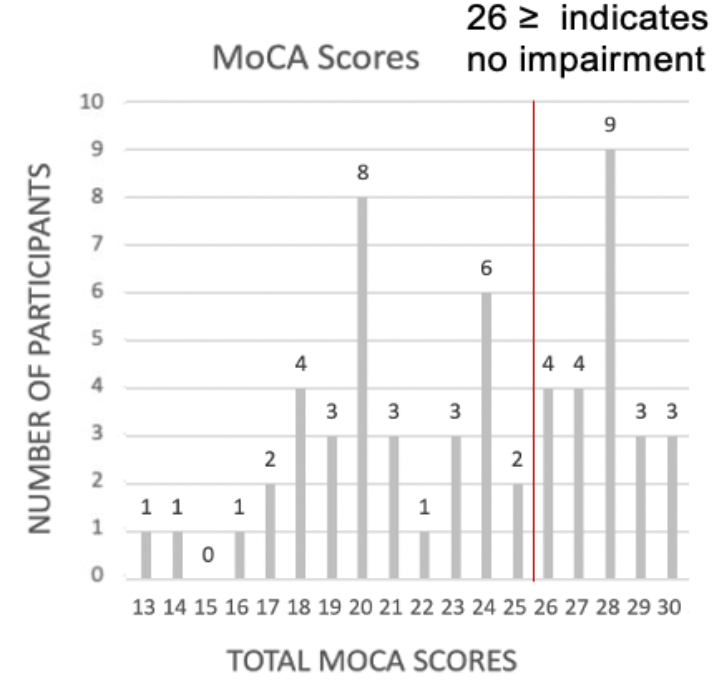
Histogram of total MoCA scores for the 58 patients who have completed the full test. Scores below 26 reflects some level of cognitive impairment.

To identify specific areas of impairment for patients post Covid-19, we divided our sample of participants who completed the MoCA in its entirety (N=58) into two groups based on their total MoCA score. Individuals in the “higher functioning” group had total MoCA scores of 26 or greater, indicating no cognitive impairment (N=17). Individuals in the “lower functioning” group had total MoCA scores of less than 26, indicating cognitive impairment (N=41). These findings are summarized in Table 1 and Figure 2. Participants with cognitive impairment scored significantly lower in executive function, attention, language, and delayed free recall domains compared to participants without cognitive impairment.

**Figure 2:**
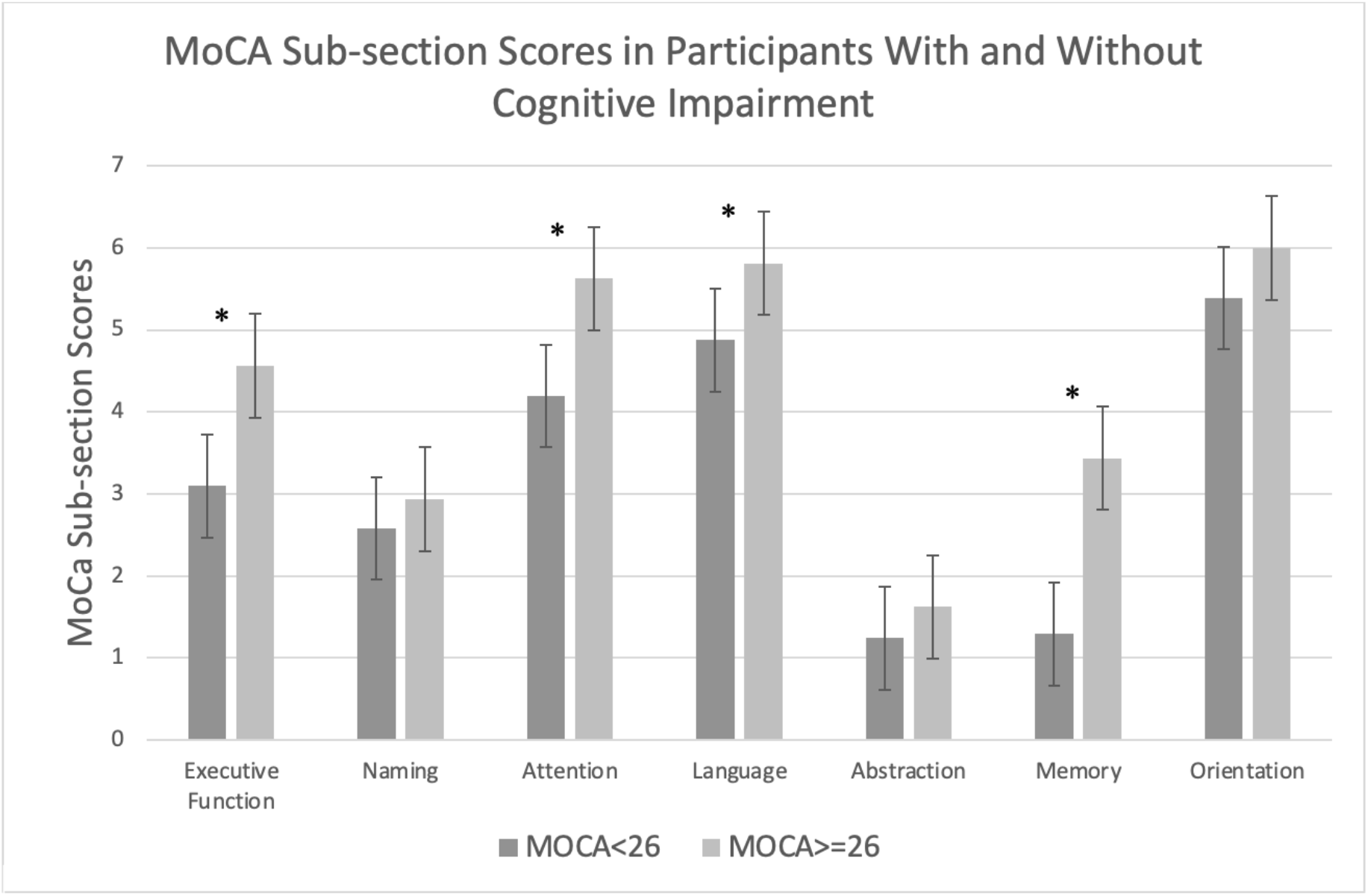
Subscore averages in participants with (total MoCA score <26) and without cognitive impairment (total MoCA score ≥26). Participants with cognitive impairment scored significantly lower in executive function, attention, language, and delayed free recall domains compared to participants without cognitive impairment.

### Rehabilitation outcomes

Overall rehabilitation scores as measured by the FIM, GG-SC, and GG-M can be found in Table 2. Average FIM scores ranged between 5.7 and 6.3, depending on the cognitive domain, indicating participants were performing tasks with supervision or partial independence. Average GG-SC and GG-M scores both fell in the 4-5 ranges, indicating they needed supervision or assistance with setup or cleanup of their tasks.

**Table 2:**
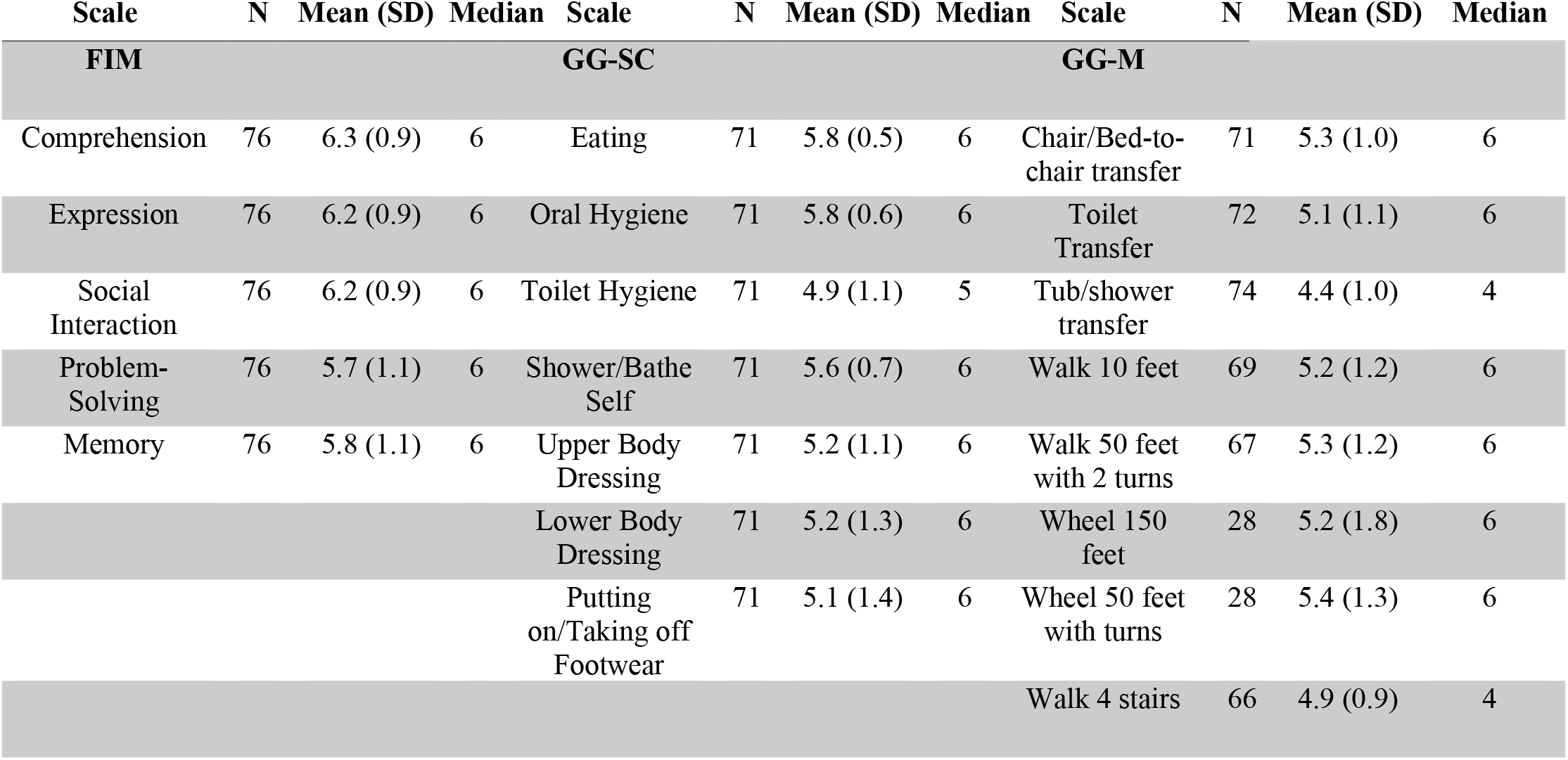
Summary of FIM, GG-SC, and GG-M scores.

#### Cognition and Rehabilitation Outcomes

Higher total MoCA scores were associated with higher FIM scores in problem solving and memory. These findings are illustrated in Figure 3 and the statistical analysis is included in Table 3. Total MoCA scores were not associated with duration of rehabilitation or GG-SC or GG-M scores (p>0.01).

**Table 3:** Spearman correlations, p-values, and Bayesian factors provided for statistically significant (in bold) and insignificant correlations between MoCA total and subscores and rehabilitation outcome measures.

|  | **Problem Solving FIM Score** | **Memory FIM Score** | **Comprehension FIM Score** | **“Walk 50 feet and turn” GG-M Score** | **“Ability to Eat” GG-SC Score** |
| --- | --- | --- | --- | --- | --- |
| **Total MoCA Score** | **$r= 0.4$ ($p=0.003$)**<br>**$BF_{10}=13.1$** | **$r= 0.4$ ($p<0.001$)**<br>**$BF_{10}=52.9$** | $r= 0.3$ ($p=0.02$),<br>$BF_{10}=6.8$ | $r= 0.2$ ($p=0.2$),<br>$BF_{10}=0.4$ | $r= 0.1$ ($p=0.5$),<br>$BF_{10}=0.2$ |
| **Executive Function** | **$r=0.4$ ($p=0.005$)**<br>**$BF_{10}=2.1$** | **$r= 0.4$ ($p=0.004$)**<br>**$BF_{10}= 2.8$** | $r= 0.2$ ($p=0.1$),<br>$BF_{10}=0.3$ | $r=0.1$ ($p=0.4$),<br>$BF_{10}=0.2$ | $r= 0.1$ ($p=0.4$),<br>$BF_{10}=0.2$ |
| **Attention** | **$r= 0.4$ ($p<.001$)**<br>**$BF_{10}=10.9$** | **$r= 0.4$ ($p<0.001$)**<br>**$BF_{10}=34.7$** | **$r= 0.4$ ($p<0.001$)**<br>**$BF_{10}=47.8$** | **$r= 0.3$ ($p=0.005$)**<br>**$BF_{10}=6.5$** | $r= 0.1$ ($p=0.2$),<br>$BF_{10}=0.3$ |
| **Language** | **$r=0.4$ ($p=0.003$)**<br>**$BF_{10}=36.5$** | **$r= 0.4$ ($p=0.003$)**<br>**$BF_{10}=62.2$** | **$r= 0.4$ ($p=0.001$)**<br>**$BF_{10}=880.3$** | $r= 0.2$ ($p=0.2$),<br>$BF_{10}=2.2$ | $r= 0.2$ ($p=0.09$),<br>$BF_{10}=3.8$ |
| **Delayed Free Recall** | $r= 0.2$ ($p=0.04$),<br>$BF_{10}=1.5$ | **$r=0.3$ ($p=0.004$)**<br>**$BF_{10}=13.6$** | $r=0.2$ ($p=0.04$),<br>$BF_{10}=1.3$ | $r=0.2$ ($p=0.04$),<br>$BF_{10}= 0.9$ | **$r= 0.3$ ($p=0.003$)**<br>**$BF_{10}=4.5$** |

**Figure 3:**
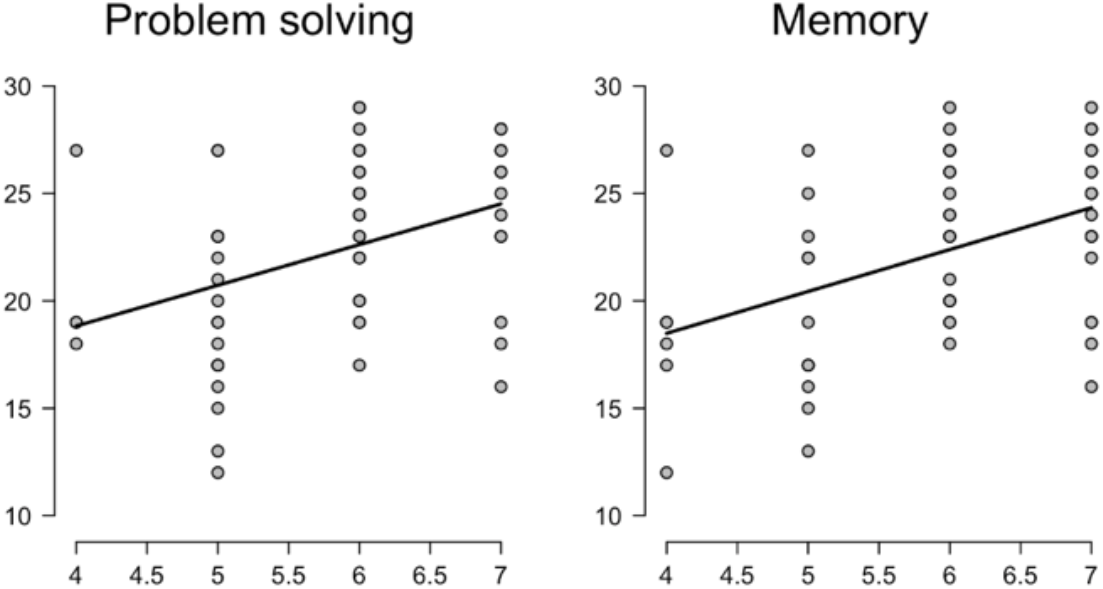
Total MoCA scores correlate positively with FIM rehabilitation scores in problem-solving and memory.

Given that executive function, attention, language, and delayed free recall were the most impaired domains in our sample, we tested how these specific subscales correlate with rehabilitation outcomes. Executive function, attention, and language MoCA subscores correlated positively with problem solving and memory FIM scores. Delayed free recall MoCA subscores correlated positively with the memory FIM score. Attention and language subscores correlated positively with comprehension FIM scores. Attention subscore positively correlated with scoring on the ‘ability to walk 50 feet and turn’ GG-M subscale. Delayed free recall MoCA subscores correlated positively with scoring on the ‘ability to eat’ GG-M subscale. MoCA subscores did not correlate with duration of stay in rehabilitation (p>0.01). The statistical findings are summarized in Table 3.

## Discussion

This is the first study to examine how cognitive status of patients in acute rehabilitation following a diagnosis of Covid-19 relates to rehabilitation outcomes. Our findings first show that majority of inpatients in acute rehabilitation after Covid-19 presented with mild to moderate cognitive impairment. Executive function, attention, language, and memory domains were the most severely impacted. These findings are in line with previous studies that reported cognitive impairment in patients with varying severity of Covid-19 disease outside of rehabilitation settings^13,27–31^. It also supports findings from two studies of patients in acute rehabilitation settings^32,33^ and expands on them by evaluating specific cognitive domains separately^33^ and by correlating cognitive function with three different rehabilitation domains: namely cognition, self-care, and motor functioning.

In line with our initial hypothesis, our data provides strong evidence that cognitive status on admission to rehabilitation predicts cognitive outcomes as measured by the FIM scale. Patients with higher overall cognitive function as well as higher scores in the executive function, attention, language, and memory subsections, achieved higher FIM scores, particularly in problem solving and memory. These findings were expected, as initial cognitive function should be a predictor of cognitive function at a future point in time.

We also found that patients with higher attention scores also scored higher in their ability to walk. Patients with higher memory scores were more capable of feeding themselves. While in both cases the correlation had strong statistical significance (p<0.01), they should be considered with caution as these specific associations were identified post hoc and were not part of our a priori hypotheses. Nonetheless, these findings are in line with previous studies of healthy older adults and people with Parkinson’s disease that found correlations between attention as well as executive function and walking ability^34,35^. Attention in addition to memory has been correlated with eating ability patients with Alzheimer’s Disease^36^. Overall, our findings suggest that screening for cognitive impairments in Covid-19 patients in rehabilitation settings can be helpful in predicting both cognitive and, to a lesser extent, self-care and motor rehabilitation outcomes.

Within the Covid-19 literature, we are currently aware of two peer-reviewed studies that evaluated cognitive functioning in the acute rehabilitation setting in Covid-19 patients. The first was a study that evaluated 87 stroke and seizure-free patients and found that 80% of them presented with cognitive deficits, especially among elderly patients^32^. The second study evaluated the cognitive profile of Covid-19 patients requiring rehabilitation and found deficits specifically within executive function, attention, and memory domains^30^. The findings in this paper are consistent with the cognitive function among participants in this study. However, neither paper examined how to cognitive profiles of their participants related to their rehabilitation outcomes.

### Study Limitations

While addressing the gap regarding to association between cognitive function and rehabilitation outcome of Covid-19 patients, our study is not free of limitations. First, our sample (N=58-80, depending on the target measure) is modest in size. Therefore, our findings cannot be easily generalized. Yet, this is one of the largest samples of Covid-19 patients evaluated in an acute rehabilitation to this date, and therefore, holds theoretical and clinical significance.

Second, this study design did not include a control group, which limits our ability to argue that the findings we report here are unique to Covid-19 patients in acute rehabilitation. Nonetheless, findings from recent studies compares patients with Covid-19 and healthy control suggest that the cognitive patterns reported here are indeed unique to this population ^3,28,29^. Moreover, while our findings are like those found in stroke patients, they differ from those found in other rehabilitation populations, including patients who suffered acute coronary syndrome (ACS). Compared to our population, patients with ACS demonstrate cognitive impairments to a lesser extent and only in the language and memory domains, not in executive function or attention^37–39^.

Third, the evaluation of cognition in this study is not sensitive to mild cognitive changes. While the MoCA is commonly used in clinical and research settings, it is a screening tool for cognitive impairment and not a diagnostic neuropsychological evaluation^40^. A more nuanced evaluation of the neuropsychological profile of Covid-19 patients can reveal additional, more subtle deficits that are not reported here.

We are also unable to causally link the cognitive deficits observed in our patient population directly to the Covid-19 diagnosis. It is possible that the lower cognitive status we reported is related to post-intensive care syndrome (PICS). PICS causes impairments in neurological, psychological, and motor function and has been found in patients who have survived serious illness requiring critical care ^41,42^. Symptoms of PICS include impairments in thinking and judgement and signs of depression and anxiety as well as muscle weakness and post-traumatic stress disorder ^43^. Clearly, there is overlap in the symptoms of Covid-19 and PICS which we cannot untangle in this study. Therefore, it is difficult to determine how much each factor contributes to disease outcome.

### Future Directions

Future studies are needed to further examine factors contributing to rehabilitation outcomes in patients with Covid-19 disease and ultimately determine how we can therapeutically target these factors to maximize outcomes. This may include components of cognitive function that can be identified with more robust neuropsychological evaluation. In addition, studies should be conducted to evaluate other measures of rehabilitation outcomes, including cardiac outcomes, pulmonary outcomes, and level of overall independence.

Long-term cognitive and mood outcomes of Covid-19 disease also need to be investigated. Our study evaluated cognition and rehabilitation outcomes shortly following disease onset and hospitalization, but it is becoming increasingly more apparent that long-term effects of Covid-19 can last months and even years^44–46^. While we are at the point where we may begin to examine 12 and 24-month sequelae of this disease, future studies are also needed to investigate lifetime impacts of severe Covid-19 several years following initial disease onset.

## Conclusion

Patients with severe Covid-19 disease requiring rehabilitation were found to have significant cognitive impairments with overall MoCA scores falling in the mild cognitive impairment category. Higher cognitive function in general and higher executive function, attention, language, and memory functions were associated with better rehabilitation outcomes, specifically in learning, memory, and some motor functioning. The findings in this study advance our understanding of the cognitive and affective attributes of patients with severe Covid-19 disease. Our study paves the way for future studies to further evaluate the associations between cognitive status and rehabilitation outcomes in inpatients post Covid-19, expecially in motor and self-care domains. By improving our understanding, the ultimate goal is to translate our findings into clinical therapies that target cognition in the rehabilitation setting to improve disease outcome.

## Data Availability

All data produced in the present study are available upon reasonable request to the authors

## Abbreviations

MoCA=: Montreal Cognitive Assessment
FIM=: Functional Index Measure
BIS=: Barthel Index Score
GG-SC=: Section GG items for measuring aspects of self-care in rehabilitation
GG-M=: Section GG items for measuring mobility in rehabilitation

## Acknowledgment

This research was supported by the Shirley Ryan Ability accelerator grant. The funders played no role in the design of this study or the interpretation of its results.

## Notes

This material has not been presented previously and there are no conflicts of interest.

### Competing Interest Statement

The authors have declared no competing interest.

### Author Declarations

Northwestern University's Institutional Review Board granted approval for our study.

## References

1. Johns Hopkins University & Medicine Coronavirus Resource Center. https://coronavirus.jhu.edu/map.html.

2. Wade DT. Rehabilitation after COVID-19: an evidence-based approach. Clin Med 2020; 20: 359–365.

3. Rogers-Brown JS, Wanga V, Okoro C, et al. Outcomes Among Patients Referred to Outpatient Rehabilitation Clinics After COVID-19 diagnosis - United States, January 2020-March 2021. MMWR Morb Mortal Wkly Rep 2021; 70: 967–971.

4. Tang D, Comish P, Kang R. The hallmarks of COVID-19 disease. PLOS Pathog 2020; 16: e1008536.

5. Rodriguez-Morales AJ, Cardona-Ospina JA, Gutiérrez-Ocampo E, et al. Clinical, laboratory and imaging features of COVID-19: A systematic review and meta-analysis. Travel Med Infect Dis 2020; 34: 101623.

6. Fauci AS, Lane HC, Redfield RR. Covid-19—navigating the uncharted. New England Journal of Medicine 2020; 382: 1268–1269.

7. Koralnik IJ, Tyler KL. COVID-19: A Global Threat to the Nervous System. Ann Neurol 2020; 88: 1–11.

8. Varatharaj A, Thomas N, Ellul MA, et al. Neurological and neuropsychiatric complications of COVID-19 in 153 patients: a UK-wide surveillance study. The Lancet Psychiatry 2020; 7: 875–882.

9. Mao L, Wang M, Chen S, et al. Neurological manifestations of hospitalized patients with COVID-19 in Wuhan, China: a retrospective case series study. MedRxiv.

10. Almeria M, Cejudo JC, Sotoca J, et al. Cognitive profile following COVID-19 infection: Clinical predictors leading to neuropsychological impairment. Brain, Behav Immun - Heal 2020; 9: 100163.

11. Hampshire A, Trender W, Chamberlain SR, et al. Cognitive deficits in people who have recovered from COVID-19 relative to controls: An N=84,285 online study. medRxiv 2020; 2020.10.20.20215863.

12. Del Brutto OH, Wu S, Mera RM, et al. Cognitive decline among individuals with history of mild symptomatic SARS-CoV-2 infection: A longitudinal prospective study nested to a population cohort. Eur J Neurol; n/a. Epub ahead of print 11 February 2021. DOI: https://doi.org/10.1111/ene.14775.

13. Becker JH, Lin JJ, Doernberg M, et al. Assessment of Cognitive Function in Patients After COVID-19 Infection. JAMA Netw Open 2021; 4: e2130645.

14. Heruti RJ, Lusky A, Dankner R, et al. Rehabilitation outcome of elderly patients after a first stroke: Effect of cognitive status at admission on the functional outcome. Arch Phys Med Rehabil 2002; 83: 742–749.

15. Mysiw WJ, Beegan JG, Gatens PF. Prospective cognitive assessment of stroke patients before inpatient rehabilitation. The relationship of the Neurobehavioral Cognitive Status Examination to functional improvement. Am J Phys Med & Rehabil 1989; 68: 168–171.

16. Yu STS, Brown T, Yu M-L, et al. Association between Older Adults’ Estimated Length of Hospital Stay and Cognitive Performance. Phys Occup Ther Geriatr 2018; 36: 136–155.

17. Nasreddine ZS, Phillips NA, Bédirian V, et al. The Montreal Cognitive ?Assessment, MoCA: A Brief Screening Tool For Mild Cognitive Impairment. J Am Geriatr Soc 2005; 53: 695–699.

18. Lam B, Middleton LE, Masellis M, et al. Criterion and Convergent Validity of the Montreal Cognitive Assessment with Screening and Standardized Neuropsychological Testing. J Am Geriatr Soc 2013; 61: 2181–2185.

19. Fiedler RC, Granger C V, Russell CF. Uniform Data System for Medical Rehabilitation: report of first admissions for 1997. Am J Phys Med Rehabil 1998; 77: 444–450.

20. Li C-Y, Graham J, Kuo Y-F, et al. Response Patterns in Standardized Functional Assessment: Section GG in Self-Care and Mobility. Arch Phys Med Rehabil 2020; 101: e12.

21. Nuzzo R. Scientific method: statistical errors. Nature 2014; 506: 150–152.

22. Wasserstein RL, Lazar NA. The ASA Statement on p-Values: Context, Process, and Purpose. Am Stat 2016; 70: 129–133.

23. Kass RE, Raftery AE. Bayes Factors. J Am Stat Assoc 1995; 90: 773–795.

24. Wagenmakers E, Verhagen J, Ly A, et al. The need for Bayesian hypothesis testing in psychological science. Psychol Sci under Scrut Recent challenges Propos Solut 2017; 123–138.

25. Dunson DB. Commentary: Practical Advantages of Bayesian Analysis of Epidemiologic Data. Am J Epidemiol 2001; 153: 1222–1226.

26. Kruschke JK. Doing bayesian data analysis: a tutorial with R, JAGS, and Stan. Second. San Diego, CA: Academic Press, 2015. Epub ahead of print 2015. DOI: https://doi.org/10.1016/B978-0-12-405888-0.09999-2.

27. Almeria M, Cejudo JC, Sotoca J, et al. Cognitive profile following COVID-19 infection: Clinical predictors leading to neuropsychological impairment. Brain, Behav immunity-health 2020; 9: 100163.

28. Woo MS, Malsy J, Pöttgen J, et al. Frequent neurocognitive deficits after recovery from mild COVID-19. Brain Commun; 2. Epub ahead of print 1 July 2020. DOI: 10.1093/braincomms/fcaa205.

29. Zhou H, Lu S, Chen J, et al. The landscape of cognitive function in recovered COVID-19 patients. J Psychiatr Res 2020; 129: 98–102.

30. Jaywant A, Vanderlind WM, Alexopoulos GS, et al. Frequency and profile of objective cognitive deficits in hospitalized patients recovering from COVID-19. Neuropsychopharmacology 2021; 46: 2235–2240.

31. Hosp JA, Dressing A, Blazhenets G, et al. Cognitive impairment and altered cerebral glucose metabolism in the subacute stage of COVID-19. Brain. Epub ahead of print 3 April 2021. DOI: 10.1093/brain/awab009.

32. Alemanno F, Houdayer E, Parma A, et al. COVID-19 cognitive deficits after respiratory assistance in the subacute phase: A COVID-rehabilitation unit experience. PLoS One 2021; 16: e0246590.

33. Patel R, Savrides I, Cahalan C, et al. COVID-19 and Cognitive Impairment: Severity, Evolution, and Functional Impact during Inpatient Rehabilitation. medRxiv 2021; 2021.03.15.21253637.

34. Gale CR, Allerhand M, Sayer AA, et al. The dynamic relationship between cognitive function and walking speed: the English Longitudinal Study of Ageing. Age (Omaha) 2014; 36: 9682.

35. Weiss A, Herman T, Giladi N, et al. Association between Community Ambulation Walking Patterns and Cognitive Function in Patients with Parkinson’s Disease: Further Insights into Motor-Cognitive Links. Park Dis 2015; 2015: 547065.

36. Hall JR, Vo HT, Johnson LA, et al. The Link between Cognitive Measures and ADLs and IADL Functioning in Mild Alzheimer’s: What Has Gender Got to Do with It? Int J Alzheimer’s Dis 2011; 2011: 276734.

37. Zhao E, Lowres N, Woolaston A, et al. Prevalence and patterns of cognitive impairment in acute coronary syndrome patients: A systematic review. Eur J Prev Cardiol 2020; 27: 284–293.

38. Silva M, Pereira E, Rocha A, et al. Neurocognitive impairment after acute coronary syndrome: Prevalence and characterization in a hospital-based cardiac rehabilitation program sample. J Cardiovasc Thorac Res 2018; 10: 70–75.

39. Salzwedel A, Heidler M-D, Haubold K, et al. Prevalence of mild cognitive impairment in employable patients after acute coronary event in cardiac rehabilitation. Vasc Health Risk Manag 2017; 13: 55–60.

40. Coen RF, Robertson DA, Kenny RA, et al. Strengths and Limitations of the MoCA for Assessing Cognitive Functioning: Findings From a Large Representative Sample of Irish Older Adults. J Geriatr Psychiatry Neurol 2016; 29: 18–24.

41. Colbenson GA, Johnson A, Wilson ME. Post-intensive care syndrome: impact, prevention, and management. Breathe (Sheffield, England) 2019; 15: 98–101.

42. Modrykamien AM. The ICU follow-up clinic: A new paradigm for intensivists. Respir Care 2012; 57: 764–772.

43. Rawal G, Yadav S, Kumar R. Post-intensive Care Syndrome: an Overview. J Transl Intern Med 2017; 5: 90–92.

44. Lopez-Leon S, Wegman-Ostrosky T, Perelman C, et al. More than 50 Long-term effects of COVID-19: a systematic review and meta-analysis. medRxiv Prepr Serv Heal Sci 2021; 2021.01.27.21250617.

45. Davis HE, Assaf GS, McCorkell L, et al. Characterizing long COVID in an international cohort: 7 months of symptoms and their impact. Available SSRN 3820561.

46. Crook H, Raza S, Nowell J, et al. Long covid—mechanisms, risk factors, and management. BMJ 2021; 374: 1648.

